# Pilot study of a ketogenic diet in bipolar disorder: a process evaluation

**DOI:** 10.1101/2024.05.15.24307102

**Authors:** Benjamin P. Rigby, Nicole Needham, Helen Grossi, Ivana Kamenska, Iain H. Campbell, Ben Meadowcroft, Frances Creasy, Cheryl Fisher, Pankaj Bahuguna, John Norrie, Gerard Thompson, Melissa C. Gibbs, Maja Mitchell-Grigorjeva, Ailsa McLellan, Tessa Moses, Karl Burgess, Rachel Brown, Michael J. Thrippleton, Harry Campbell, Daniel J. Smith, Sharon A. Simpson

## Abstract

**Background:** Bipolar disorder is a serious mental illness, which requires new strategies for prevention and management. Recent evidence suggests that a ketogenic diet may be an effective intervention. This research aimed to explore the feasibility and acceptability of a ketogenic diet intervention for bipolar disorder, fidelity to its behavioural components and the experiences of the participants and research clinicians involved.

**Methods:** A mixed-methods process evaluation was conducted. Semi-structured telephone interviews were carried out with 15 participants 1-2 months after completing a 6-8 week modified ketogenic diet intervention, and 4 research clinicians from the study team following the completion of data collection. Data were thematically analysed. Fidelity checklists completed by research dietitians were analysed using descriptive count and percentage statistics. Findings are reported post-hoc, following the analysis and publication of the main pilot study findings.

**Results:** Qualitative data indicated that participants had various motives for taking part in the study, including weight loss. It was important to support people’s motives while facilitating clear and realistic expectations. Despite the challenges of initiating and maintaining a ketogenic diet, including for some its disruptive effects on daily living, many participants perceived physical and psychological benefits (e.g. significant weight loss, mood stability and an enhanced ability to focus). A range of behavioural (*e.g.* goal setting), social (*e.g.* family and dietitians) and technological (*e.g.* apps for monitoring) support mechanisms were generally considered key facilitating factors. Meanwhile, dietary preferences, concerns about the diet and its impact, the testing burden and capacity of the delivery team were perceived as barriers for some. The importance of wider contextual influences (*e.g.* the cost of living and sociocultural expectations) were highlighted. Overall, descriptive analyses indicated moderate-to-good fidelity to the behaviour change components of the study.

**Conclusion:** We provide novel insight into the experiences of people living with bipolar disorder initiating and following a ketogenic diet, as well as those of research clinicians who support the intervention. Future trials may benefit from increased clinical research capacity, better-defined entry and exit routes, additional interpersonal support, and greater understanding of how social and societal factors impact participation.

**Trial registration:** Study registration number: ISRCTN6163198 (02 March 2022)

## Background

Bipolar disorder (types I and II) is a serious mental illness characterised by episodes of elevated mood and depression [1]. It has a lifetime risk of 1-2% [2] and is considered among the most challenging psychiatric disorders to manage [1]. Typically presenting during early adulthood, bipolar disorder is associated with significant economic and societal impact, comorbidities and high mortality [3, 4]. Life expectancy among people with this disorder may be up to 20 years less than the general population, with suicide and cardiovascular disease being leading contributors to excess deaths [1, 3]. However, current strategies for prevention and management of bipolar disorder are suboptimal and new interventions are required.

The ketogenic diet, premised on very low carbohydrate and very high fat intake [5], represents an intervention with emerging therapeutic potential across a range of psychiatric conditions, including bipolar disorder [6–9]. It is proposed that this diet may contribute to more stable energy production in the brain through the metabolism of ketones as an alternative energy source to glucose [10], thus overcoming issues of glucose metabolism and insulin resistance that are common among people with bipolar disorder [11–14]. However, despite promising case reports, there remain no randomised controlled trials (RCTs) examining the effect of a ketogenic diet in bipolar disorder.

A recent four-month pilot trial investigated the effects of a modified ketogenic diet on individuals with schizophrenia or bipolar disorder with existing metabolic abnormalities [9]. Among participants adherent to the diet, metabolic improvements were reported (*e.g.* reduced BMI and waist circumference measurements), as well as enhanced life satisfaction and sleep. In another recent pilot study, Needham *et al.* investigated the recruitment, feasibility and acceptability of a modified 6-8 week ketogenic diet in bipolar disorder among euthymic participants (*i.e.* those in a period of emotional homeostasis) [4, 5]. The key findings were that intervention recruitment and retention were feasible, and that most participants reached and maintained ketosis during the study period. Combined with promising outcomes data from this study (publication forthcoming [15]), these findings suggested that a full clinical trial is warranted [4, 15]. To complement this latter pilot study and inform the development of a future RCT (*e.g.* through understanding factors that influence study design or participant adherence to intervention), we conducted a mixed-methods process evaluation of the pilot post-intervention.

Process evaluations differ to outcome-focused research by exploring how interventions work in practice through increasing understanding of the implementation of interventions, the mechanisms of impact and the context that shapes these two factors [16]. Both quantitative and qualitative analyses can contribute to these aims [16, 17]. In the piloting phase, process evaluation helps understand fidelity, feasibility and acceptability, and optimise the future design and evaluation of full-scale trials [16]. Such considerations are particularly important in developing trials for population groups with severe mental illness, such as bipolar disorder, who may have complex barriers to research involvement [18]. For example, people with bipolar disorder are at risk of therapeutic misconception or optimistic bias when presented with research information [19], and often experience impaired concentration or memory [20]. This may affect the acceptability of and adherence to interventions such as the ketogenic diet, which is known to challenging albeit achievable among adults [21–23]. Previous research has indicated that numerous components may be important to support adherence to and outcomes from dietary interventions for people with bipolar disorder, including goal setting, regular and tailored support from healthcare and allied professionals, and psychological therapies [24]. However, no studies have examined these and associated factors in relation to the ketogenic diet as a therapeutic intervention for bipolar disorder.

As part of a wider pilot trial [4, 15], this research aimed to further explore the feasibility and acceptability of the ketogenic diet intervention and elements of the study design, examine fidelity to the behavioural components of the intervention, and explore the experiences of participants and research clinicians involved in the research.

## Methods

### The pilot study

A favourable ethical opinion was obtained for this study by the South East Scotland Research Ethics Committee 02 (approval number: 22/SS/0007) and it was approved by NHS Lothian Research and Development. The primary aim of the pilot study was to assess recruitment, acceptability and feasibility of a 6-8 week trial of a modified ketogenic diet for individuals with bipolar disorder. Secondary objectives were to assess the relationships of a biomechanical, metabolomic and brain imaging biomarkers with clinical and functional outcomes. In brief, the intervention comprised a diet with estimated energy requirements of approximately 60-75% from fat, 5-7% from carbohydrate and additional calories from protein [5]. Participants received tailored support from an experienced ketogenic dietitian, and had access to a psychiatrist on the research team if required. The support package was informed by the COM-B framework [25] and behaviour change theories [26, 27]. Twenty-seven euthymic individuals were recruited into the study. Full details about the study design, intervention components and participants are published elsewhere [4]. The clinical and functional outcomes will also be presented in a forthcoming publication [15]. The remainder of this paper reports on sampling, recruitment, procedures and findings for this additional process evaluation component of the study.

### Participants

Fifteen of the 27 individuals recruited into the pilot study were purposively sampled and agreed to be interviewed. We aimed to achieve variation in participants’ age, gender, socioeconomic background and whether they completed the 6-8 week intervention. The Scottish Index of Multiple Deprivation (SIMD) was used as proxy measure of socioeconomic background, recording participants’ residential status on a scale of 1^st^ decile (most deprived) to 10^th^ decile (least deprived). Participant characteristics are reported in Table 1.

**Table 1.** Participant characteristics (pilot study: n = 15)

| Characteristics |  |
| --- | --- |
| Age | Mean = 48.5 years (range 37-54) |
| Sex | Female = 10<br>Male = 5 |
| Place of residence (SIMD) | 1<sup>st</sup> and 2<sup>nd</sup> deciles = 0<br>3<sup>rd</sup> and 4<sup>th</sup> deciles = 4<br>5<sup>th</sup> and 6<sup>th</sup> deciles = 1<br>7<sup>th</sup> and 8<sup>th</sup> deciles = 3<br>9<sup>th</sup> and 10<sup>th</sup> deciles = 7 |

Table 1 shows that two-thirds of the participants were female and two-thirds were resident in areas in the four least deprived SIMD deciles. One of the 15 discontinued the diet before the 6-8 week intervention periods was completed. Four research clinicians (all female) involved in the delivery of the intervention were also purposively recruited and agreed to be interviewed: two research psychiatrists (one of whom also acted as study coordinator) and two research dietitians. The descriptive quantitative analysis of the fidelity checklist comprised data from the 25 participants for whom at least one diet review meeting was recorded.

### Procedure

#### Qualitative component

One-to-one semi-structured telephone interviews were conducted with study participants 1-2 months after completing the 6-8 week diet intervention, and research clinicians implementing the intervention following the completion of the intervention component of the study. The use of a semi-structured interview format allowed flexibility to ask additional questions based on the participants’ responses, and a set of prompt questions were used to stimulate discussion if needed [28]. Prior to the interviews, all participants were asked to read an information sheet and provide informed consent. All interviews were conducted by BPR, recorded on an encrypted electronic device, transcribed intelligent verbatim and the transcripts de-identified. Interviews with the pilot participants lasted between 37 and 98 minutes. They were structured around the following topics: recruitment into the study; experiences of being a research participant; the diet; the support package; perceived outcomes; and areas for suggested development. Additional file 1 provides the interview guide. Interviews with research clinicians lasted between 22 and 45 minutes, and were structured around: general experiences; implementing the study; engagement with participants; and future directions (see Additional file 2 for the interview guide).

#### Quantitative component

Data were taken from the 104 diet review meetings in which fidelity checklists were completed in conversation between participants and a research dietitian. The checklists (see Additional file 3) were used by dietitians to ensure coverage of key behavioural strategies that underpinned the intervention: i) providing information on the benefits of changing diet; ii) goal setting and creating an action plan; iii) social support; iv) relapse prevention strategies; v) reviewing the outcome of goals and plans; vi) discussing (potential) barriers and how to overcome them; vii) self-monitoring/feedback; viii) discussing performance and providing feedback; ix) discussing environmental restructuring; x) discussing how doing well could be rewarded. Hand-written checklists were compiled and cleaned by HG, IK and NN, then converted into digital MS Excel records by BPR.

### Analysis

Analyses were initially performed by BPR both alongside and after the examination of study outcomes; findings are reported post-hoc [16]. Qualitative data were analysed using an inductive thematic approach based on Braun and Clarke’s original six-step method [29]. SAS reviewed two transcripts independently and interpretation of the data were discussed between BPR and SAS, as well as in team meetings. Quantitative data were initially analysed by BPR using descriptive counts and percentages to examine the proportion of meetings in which each behavioural strategy was covered and patterns in their use. Two sets of findings were generated. First, an overall assessment of fidelity that included all 25 participants with completed checklists. Second, an adjusted assessment that only included data from participants that completed the diet at 6-8 weeks follow-up (n=20). Data were not included in either analysis from any of the participants’ first meetings for two components, which by definition were not relevant to be discussed at that time point (reviewing goals and plans; and discussing performance). The interpretation of data were discussed at research team meetings.

## Results and discussion

### Qualitative findings

Five salient themes were identified in the qualitative data relating to recruitment, feasibility, acceptability and implementation of the ketogenic diet intervention. A summary of themes and their associated sub-themes is provided in Table 2.

**Table 2.** Summary of themes and sub-themes.

| Main theme | Subthemes |
| --- | --- |
| Encouraging entry and supporting exit | Addressing participant motives |
|  | Setting clear expectations |
|  | Providing exit routes |
| Challenging but potentially transformational | A big undertaking |
|  | Perceived impact |
| Intervention facilitators | Behavioural support mechanisms |
|  | Interpersonal support |
|  | Teamwork |
|  | Technology |
| Intervention barriers | The testing burden |
|  | Capacity of the delivery team |
|  | Dietary preferences |
|  | Participants' concerns |
| The wider context | Social pressures |
|  | Keto in a cost-of-living crisis |

#### Theme 1 – Encouraging entry and supporting exit

Findings from the pilot study suggested that recruitment and retention of participants was feasible [4]. This was reflected in the majority of experiences articulated through the interviews. While recruitment to this pilot was conducted primarily through Bipolar Scotland, the underlying mechanisms that encouraged entry into the study may be applicable in alternative contexts. For example, providing study-related resources so that people are *‘able to learn quite a lot about it before’* participation, through the website, *‘factsheets, and information about the study* (Participant 7)*.’* Further, some participants valued hearing the experiences of a member of the research team who has a diagnosis of bipolar disorder and follows a ketogenic diet.

He spoke, his presentation was excellent. And he lives with bipolar and he’s been on the ketogenic diet for about eight years and he could talk to the benefits that he felt. So it was very powerful (Participant 15). Given that self-efficacy among people with bipolar disorder may typically be low [30], such testimonies have the potential to enhance people’s belief in their abilities through encouragement and vicarious experiences [26], thus supporting them to engage with a novel and challenging activity, such as following a new diet. This approach may also support motivation through feelings of relatedness [31], and help set clear expectations of the study.

##### Addressing participant motives

Participants entered the pilot study with various experiences of living with bipolar disorder. Some had been diagnosed many years ago, others as little as six-to-nine months prior to recruitment. Their ongoing treatment and management varied, as did their motives for participating. While participants sought alleviation of their symptoms, four wider and potentially stronger motives were identified. For some, the study represented hope. This was unsurprising given the challenge of managing this severe mental illness [1, 4].

> I hoped that the keto diet would do something I haven’t been able to do on my own…I’d hoped that the keto diet would be a miraculous cure. I was looking for help or support to try and help me get better (Participant 8).

Other participants cited a desire to reduce drug-use and challenge *‘the current medical model for someone who has a diagnosis of bipolar [which] is to take medications and lower your expectations* (Participant 5)*.’* Altruism was a third motive identified in the data.

> I decided to participate because I felt like it was a very worthwhile thing to do…for other people and in the future. And for the knowledge that will be obtained from it, they’re actually treating people (Participant 1).

Eighteen of the 27 participants in the pilot study were overweight or obese [4], which is common among people with bipolar disorder [32]. For many, weight loss was a primary motive.

> The anti-psychotic drugs make me crave sugar and my weight has just yo-yoed up and down. And I know that I should be eating low carb, it seems to work for me, it seems to really work for the weight loss. However, any time I’ve tried it before it just sends my mood loopy. I take more anti-psychotics, which makes me crave the sugar, and then it just goes round and round…as soon as I saw it, I was like, “I want to take part in this (Participant 13).”

In future trials, it will be important to ensure that communications and recruitment strategies support a range of potential motives, which may extend beyond direct presentations of bipolar disorder. While significant weight loss has been observed in this pilot study and others [9, 15], the weight loss motive, as well as hope, should be addressed cautiously, given both the absence of high-quality RCT evidence of the effectiveness of interventions used in the management of obesity in people with bipolar disorder [32], and the risk of therapeutic misconception or optimistic bias in this population subgroup [19]. This reinforces the need for clear expectations around involvement in studies.

##### Setting clear expectations

Given how challenging it can be to follow a ketogenic diet [5], it is important that potential participants are fully informed about the reality of participating in such a study. Several participants felt that the expectations were clear and realistic, which seemed in some cases to be linked to past experiences of medical intervention or research.

> I’d been told before the appointments that’ll last this length of time…And I knew in advance about the once a week phone calls. And taking the blood samples and texting the results. Nothing took me by surprise…blood tests, I’ve had plenty of them. I took part in some research when I was first psychotic and even had the full brain scan at that point (Participant 13).

Meanwhile, others felt that expectations around the diet and testing regimens could be made clearer prior to engagement.

> And I have to say, certain things, particularly in the first week, really threw me. I had no idea that’s what I was going to be involved in (Participant 15).

These experiences led the research clinicians to reflect and adjust their practice as the pilot continued. However, they also noted that baseline expectations were very high and may need to be managed carefully, as noted in previous research among people with bipolar disorder [33].

Furthermore, in pharmaceutical contexts, expectations of bipolar disorder treatment can significantly influence adherence to behaviours [34], and such factors should not be discounted in behavioural interventions.

> I think we have discussed a bit as a team, and I think it would be really helpful to spend a little bit more time with people before they started the study to make sure that they were really clear exactly what the study would entail (Research Psychiatrist 1).

> It was a different group with different needs, with different expectations, and the expectations, I would say, of the participants was extremely high. Because there’s so very little in terms of what can actually be done for bipolar disorder…the fact that looking to use this diet, I would say that the expectations of the participants were very high. Maybe that’s something that we need to be very aware of (Research Dietitian 1).

##### Providing exit routes

During the pilot, 77% (20 of 26) participants remained on the diet until 6-8 week follow-up [4]. One-third were reported to maintain their diet post cessation of the study period [15]. When interviewed one-to-two months post-follow-up, seven of 15 participants either stated that they remained on the diet, or expressed a desire to try the diet again. However, in most cases they reported concerns about the loss of support that came with being part of a research study, which may be compounded by the complexity of the ketogenic diet [5].

> It’s not a simple thing and I was doing quite a lot of calculations and all that kind of stuff. And I suppose coming to the end of it and trying to find out how I can continue…because it really helped me, I felt so much better…I’m trying to find a way to do it. I’m not sure any of these ways [available online] are going to be as effective as the medical version, but I’ll keep experimenting. I think my understanding was that…staying on the version that I was on with the study isn’t advisable without supervision (Participant 6).

One of the research clinicians also reflected on the need for robust clinical protocols to support cessation or withdrawal from the dietary intervention.

> I wasn’t maybe exactly sure how to explain to people how the withdrawal process worked or what next…and as you said, providing any aftercare reports. Some people just completely disengaged, so it was really difficult, we couldn’t even get in contact with them after they’d made that decision. And I suppose that’s another thing, should there be a process in place as to what to do if somebody completely disengages (Research Psychiatrist 1)?

Anecdotal evidence suggests that trial participants experience various emotions when their involvement in a study ends, with isolation and uncertainty among other feelings being common [35]. Recognising this is particularly important for studies involving people with bipolar disorder, who may experience greater social isolation and loneliness compared to the general population [36, 37]. Findings from this study highlight the need to support participants’ motives both pre- and post-intervention, which includes setting clear expectations. This will enable participants to appreciate the potential challenges and rewards associated with interventions such as the ketogenic diet.

#### Theme 2 – Challenging but potentially transformational

In general, participants felt that being on the ketogenic diet was challenging. However, many of those interviewed considered the undertaking to be acceptable given the perceived benefits. For some, this was a transformational lived experience.

##### A big undertaking

Transitioning to and maintaining a ketogenic diet can be challenging for adults [21–23]. In this study, some participants recognised the value of being *‘recommended that I staged the introduction of ketogenic meals* (Participant 2),*’* and there is some evidence to support this from other clinical settings, which suggests better toleration [38]. For many, maintaining a daily blood ketone testing regimen, shopping and preparing food were perceived to be *‘very time consuming* (Participant 10)*’* and take considerable effort and preparation, particularly in the early stages of the intervention period, thus reflecting previous research.

> It was quite tricky at first, but I quickly realised what was making it tricky was cooking for the other four [in the home]. And at first, I was like, I’ll try and incorporate meals that we can all eat. And then I was like no, that’s not going to work…So it was kind of like having to think about two lots of eating, what I was eating and what they were eating. But that got easier (Participant 19).

For several participants, this balancing of the diet with family commitments, including meal times, was just one of several ways in which the intervention was considered disruptive to everyday activities. Often, these perceptions related to the social aspect of eating.

> Like how am I going to socialise with food so much a part of that in our culture? So I hadn’t somehow realised that that could be a barrier. And then carrying around the things that I needed with me if I was going somewhere. I decided not to go anywhere for the first couple of weeks to keep it easier. I think it might have been helpful to just know that, maybe you put this in place at a point where you can just take your time whilst you get into ketosis (Participant 1).

For a minority of participants, this sense of challenge may have been exacerbated due to cognitive difficulties.

> I think probably the main barrier was lithium because measuring things and then going back to the charts…When I started having to calculate things, and I don’t necessarily think it would be difficult for an ordinary person, to be honest, but for me it was like, oh my God, this is like a major mental meltdown situation (Participant 14).

Impaired concentration and memory are common among people with bipolar disorder [20].

However, similar symptoms may manifest with the onset of ‘keto flu’, which is a cluster of transient symptoms that can be reported as occurring within the early weeks of a ketogenic diet [39]. While it is not possible to discern the relative impact of these two possibilities in our study, it highlights a potential barrier.

These findings indicate that people with bipolar disorder may find the early stages of being on a ketogenic diet to be a considerable adjustment before familiarisation, particularly where family circumstances are considered. They suggest that additional support may be required in the initial few weeks, which are likely to be the most difficult [40], enabling participants to engage for long enough to experience perceived benefits.

##### Perceived impact

The main outcome findings of this study are reported elsewhere [15]. However, all participants discussed their perceptions of the study’s impact, which often corroborated quantitative observations of weight loss and reduced mood lability, as well as results in other studies [9]. Additional benefits such as self-control and discipline were reported (Additional file 4 provides a full list of benefits reported during interviews).

> I think even within the first two weeks, I felt like I got my brain back. It was an exceptionally stressful time, and it was nothing to do with the study. It just was in my life, like a lot of stress or highly stressful things to deal with. But I managed okay. I kept things to myself. I kept thinking to myself other times in my life this would’ve sent me under by now. These things would’ve sent me under, into some spiral of either high anxiety or depression (Participant 1).

> As soon as I had used the keto for two or three days, I just felt more relaxed, more peaceful than I think I’ve ever felt in my life. And in that whole period since then, I don’t think I’ve experienced any fear or anxieties, hardly any agitations. This has always been a big problem, I seem to get very angry, so clearly that’s disappeared (Participant 14).

> I’ve lost a stone and a half, probably more actually, nearer two stone. So that was a big thing for me (Participant 11).

These findings are particularly noteworthy given the difficulty of managing bipolar disorder [1], and point toward the therapeutic potential of the ketogenic diet to address both a key characterisation of this severe mental illness (i.e. changes in mood) and commonly associated cardiometabolic risk factors in this population. The potentially transformational extent of these perceived and observed benefits for some participants may help explain why they undertook and continued a significant lifestyle intervention, even after the study had finished. These perceived outcomes contributed strongly to its sense of acceptability, despite the challenges.

However, many participants perceived a range of side-effects from the intervention that, as reported elsewhere, were generally mild and usually resolved with dietary adjustments [4]. For some, the lack of noticeable improvement in their mental or physical health status could be disappointing.

> It’s likely out of frustration and that was just because I wasn’t seeing any positive changes as in weight loss, like proper changes…My mood would really dip when I did my blood tests, my finger prick and the ketones were high or the glucose…I was hypoglycaemic for a couple of times. But I got really down when I’d done everything and the ketones were high [but saw no results] (Participant 15).

Examples of negative experiences are also included in Additional file 4. To manage expectations, and overcome set-backs or side-effects, it is possible that the intensive tailored support received by participants in this pilot [4] may have triggered key mechanisms that facilitated adherence.

#### Theme 3 – Intervention facilitators

##### Behavioural support mechanisms

Previous research has tended to overlook the importance of different behaviour change techniques in dietary interventions [41]. Prompted by the intervention fidelity checklist (see Additional file 3 and quantitative findings for more detail), research clinicians supported participants to implement a range of behavioural support mechanisms tailored to participants’ needs, which they reported as being beneficial for adherence to the diet and testing regimen. These included action planning [42], behavioural prompts [43] and self-monitoring [44].

> I’ve got a reminder in my work calendar, and then I’ve got a reminder in my personal calendar, and then if I’ve got a hectic day, I’m also setting a phone reminder so that an alert goes off (Participant 5).

> I think it was just working out a good system, because we eventually got, like, a small diary, and then just wrote the recipes in it (Participant 7).

Another key behavioural strategy was setting and evaluating goals [45].

> Every week we looked at what their goals were going to be for the next week, if we made changes, what kind of adverse events were they having (Research Dietitian 1)?

While importantly goal setting primarily supported adherence to the diet [46], the intervention may have also prompted some participants to reflect or act on bigger lifestyle goals. It is suggested that interventions for people with bipolar disorder need to extend beyond a narrow focus on adherence through traditional behaviour change techniques, to consider self-worth [47] and problem-solving that may underpin longer-term behavioural change [24]. For example, *‘my diet generally was absolutely shocking, so I figured it would encourage me at least to have some clear goal, even if it was from a physical health point of view* (Participant 11)*.’* These wider goals may have been facilitated through behavioural and interpersonal support.

##### Interpersonal support

It was apparent that social support provided to participants by family, research clinicians involved in the study, and existing care teams was crucial, leaving some participants doubting the potential for the intervention to work outside the support of a research project.

> And [my spouse] just helped me out, like she said, “if you’re having a brain meltdown just grab me, I’ll fix it.” So they’re a bit of an angel so I was very lucky. For a while we ate separate meals and then we both decided we were quite lonely so we started to combine them, which was much nicer (Participant 14).

> I liked working with Research Coordinator and Research Dietitians, that was fantastic, great support from both of them…I don’t think people could really do this sensibly without that kind of support, it’s hard. And reflecting back on it. There are a few habits that I’ve gone back to (Participant 15).

This reflects research that argues providing consistent interpersonal support from family and delivery teams is pivotal for ketogenic diet interventions [48–50]. Some participants reported that close monitoring by the research clinicians gave them confidence in the study’s procedures.

> I thought the support was really good. The dietitian was phoning you all the time and if you had questions, you could contact people, so you didn’t feel alone in a particular way (Participant 3).

Participants also commented on the contact time between themselves and the research clinicians (typically at least one diet review meeting per week). Most felt that this was adequate, while for some *‘it was probably better than I thought it would be* (Participant 4)*.’* Most participants appreciated the responsiveness of the study team, their reassurance, and the way in which the diet, through the provision of recipes and resources, was tailored to their individual needs and preferences by one of the dietitians. This reflects previously observed benefits of tailored dietary information [51], despite concerns about a lack of practical examples of effective tailoring to dietary preferences in real-world weight loss contexts [52].

> I don’t have issues about the amount of fat, but I do have some concerns about the kinds of fat. So the dietitian and I have worked on looking at this particular kind of fat and maybe I could switch it out for something (Participant 5).

While social support, broadly conceived, was evidently a strong facilitatory factor for participants and should be considered as a component of a future trial, it also played an important role among research clinicians delivering the intervention.

##### Teamwork

This pilot study had numerous components and involved a large team of people drawn from different academic and healthcare backgrounds. For example, psychiatry, radiology, neurology, dietetics and behavioural sciences. It was apparent that the cohesiveness of the research intervention team contributed to the feasibility of implementation. All research clinicians interviewed commented on the strength of the teamwork they experienced.

> There were situations where I needed to contact The Study Coordinator, she responded quickly and we managed to resolve any issues or any further information…I did have someone else to direct any of the queries to and they were picked up and actioned by The Study Coordinator (Research Dietitian 2).

Underpinning this teamwork was a sense of effective communication and the use of modelling.

> It was useful having the dietitian there as well because I talked it through with The Study Coordinator in the practice session, to sort of make sure that I was comfortable. There was a bit of delay between that session and seeing my first participant. The dietitian stepped in to do it again, so I got another viewing of actually how to [support] (Research Psychiatrist 2).

This supports research that indicates implementation of complex, multispecialty clinical studies may require foundations of effective teamwork and communication strategies [53]. Shared study leadership and regular meetings between clinical and research staff, as used in this pilot, are considered important features of teamwork that can bridge and embrace different specialities [54].

##### Technology

Different forms of technology were used during the study. These included text messaging, video calling and an app. Originally, all participants were supposed to have access to two apps through devices such as their mobile phones, to support logging and transfer of data from daily capillary readings and mood scales [4]. However, due to licensing issues, only participants recruited later into the study were able to access the Ketomojo app (for capillary readings). The use of the app was welcomed by both participants and research clinicians.

> I think the app is essential for moving forward…That’s because all I did was pull up the device, draw the blood and then put it to one side and then carry on. If I had been able to log in the emotional stuff at that point it would have been good (Participant 5).

> Rather than them text, they moved their data to Ketomojo. That was easy as well. Also, because if they forgot to send their data one day or they forgot to text it, we would text them to remind them to send it. Whereas if it was on the Ketomojo app, it was already there (Research Dietitian 1).

These experiences demonstrate how the use of technological developments can support with behaviour change techniques, such as prompts [43], but also contribute to the potential acceptability and feasibility of an intervention. Future trials may wish to explore the role of technological components more fully, including developments such as continuous ketone monitoring [55, 56]. The effective use of technology in the delivery of the intervention has the potential to reduce barriers such as the administrative burden on both participants and research clinicians.

#### Theme 4 – Intervention barriers

All participants and research clinicians commented on a range of potential barriers to the intervention. Broadly, these comprised those relating to implementation and delivery of the intervention, or constraints at the individual-level for participants. These were generally factors that could be managed during the intervention period, or have acceptable solutions for the future.

##### The testing burden

As part of the pilot, participants had initial and follow up meetings with a research dietitian and psychiatrist, as well as pre- and post-intervention blood tests and MRI scans. They were also asked to measure blood ketone and glucose levels and recorded elements of their mental state on a daily basis [4]. Many commented favourably on the comprehensiveness of these processes. However, some participants considered the testing regimen (i.e. both for outcomes and as part of the intervention) to be burdensome.

> The very first [day of tests], I was exhausted. I’d come down from the north of Scotland and I didn’t quite know where I was going then. I eventually got myself there and I think there was a lot of questions. So it was quite heavy going to be honest…And then the next day I had the MRI and I didn’t sleep terribly well in the hotel. I fell asleep in the MRI (Participant 3).

This was also recognised by the research clinicians.

> It could feel quite a lot seeing two people in one day, but I think it was perfectly manageable. I suppose, thinking from the participants’ point of view, it might have been too much. I tended to see them first, and so my bit was relatively more straightforward, but I know that they then give them lots of information with Research Dietitian 1. But she gave them chance to consolidate that (Research Psychiatrist 1).

The idea of a physical and emotional burden around the testing components of the study was also evident in relation to daily blood capillary readings for ketone and glucose levels. Some found the prospect of *‘finger pricking every day for two months, quite a lot to get your head round* (Participant 1)*,’* while others felt a strong sense of responsibility to ensure that they were testing appropriately and providing good data.

> I would worry that things had been recorded properly. Especially when I had to deal with two separate apps and for me it was so important that I did this properly. Like if I was going to do it, I was going to do it 100 per cent, as best as I could, you know what I mean? (Participant 1).

Our findings reflect previous diabetes-related research that highlighted the potentially intensive and burdensome nature of daily self-monitoring of blood glucose and ketone levels [57, 58]. Several participants commented on the importance of reassurance, education and a positive baseline testing experience for their engagement and adherence. In future trials, research clinicians should adopt a flexible approach to supporting emotional reactions to testing and continue to provide individualised assistance if required [57].

> I was struggling with the Ketomojo device at the very start. I couldn’t seem to draw blood, I couldn’t seem to get the blood in the little thing, and it ended up The Study Coordinator was on the phone with me talking me through it (Participant 5).

##### Capacity of the delivery team

Three of the four research clinicians interviewed commented on difficulties they sometimes had with the study administration. Our findings highlighted ways that may support with these intervention-related tasks in future, for example the use of technology.

> The paperwork burden is quite big, so you wonder about whether that could be done in a paper-light fashion so it could just be done online (Research Dietitian 2).

However, technology is no guarantee of a reduced administrative burden in clinical trials [59]. Some other challenges faced by the research clinicians were more measurement-related.

> It was necessarily participant facing. It was just slightly tricky at points to arrange MRI scans at the research facility at the right time, because they only had fixed slots, and clearly participants have busy lives. That was quite often more difficult. Sometimes, when we were booking people in quite last minute, to find them hotels, the university service didn’t always have hotels available that quickly (Research Psychiatrist 1).

While impossible to offer a ‘walk-in’ service, these findings suggest that in any future larger trials, there needs to be sufficient capacity among the delivery team to undertake the administrative task load. Further, it may be necessary to increase the support available to participants, particularly dietetics.

> If it’s a bigger number, you will need more dietitians…I mean, just to give you an example, when I said to you that we started seven people in one month, generally, within epilepsy, you would maybe start two or three a month. So starting seven in a month was hard, but it is doable. I think it would be important to know where our study participants were coming from, because our volunteers were all well [euthymic] people with bipolar disorder (Research Dietitian 1).

##### Dietary preferences

The experienced ketogenic dietitians worked closely with participants to tailor the intervention. This input ensured that, as far as was practicable, the prescribed recipes and suggestions reduced the risk of monotony associated with the diet [60], and aligned with participants’ dietary preferences. Unsurprisingly, some participants commented that there were aspects of the diet they did not like [61].

> It was very hard to consume that amount of fat each day, that was another thing I wasn’t aware, that’s what the keto diet was all about. So, you know, you weigh out olive oil in the salad, you’re basically drinking olive oil at the end of the salad (Participant 15).

Research in paediatric groups has shown that dietary preferences have mixed impact on the ketogenic diet. For example, while existing preference for high-fat food types is not considered to influence adherence [62], overall dietary preferences are one of the strongest drivers of discontinuation [63]. There remains work to be done to better understand the role of preferences in adults with bipolar disorder, who can experience concomitant challenges of disordered eating.

##### Participants’ concerns

It was apparent that participants had concerns about their involvement in this study, with some reporting in interviews that on occasions they experienced an elevated state of arousal or anxiety.

> I think I was always a bit edgy about have I prepared. I was a bit hungry as well sometimes and it works as an appetite suppressant. I was a bit edgy about, just in general feeling that keto, keto, always talking about keto (Participant 4).

While concerns were generally perceived as mild and easily managed through self-regulation strategies, and the majority of participants experienced stable or improved mood lability, a minority of participants expressed feeling particularly anxious about the potential impact of the diet on their mood state, toward either hypomania/mania or depression, even if this did not occur.

> One of the concerns I had was going into hypomania, just because of the change in my diet, big change and how that would impact my mood (Participant 12).

> I suddenly got this anxiety that I was going to trigger a depressive episode, which I haven’t had for several years…The kids go to their dad’s about 40 per cent of the time, so I don’t have much backup (Participant 2).

At the time of entry into the study, participants were euthymic; only one participant experienced a period of hypomania and another a depressive episode during the intervention period, each lasting one month [4]. Nevertheless, these examples highlight the importance of understanding perceptions of mood among participants with bipolar disorder. In particular, while lifestyle changes have typically been studied for their therapeutic potential, little attention has been paid to how stressful or disruptive these may be. While all participants in the study had at least some contact with a psychiatrist on the study team [4], given that there is some evidence around the triggering effects of stress on depressive episodes in particular [64], it may be prudent for future studies to consider more regular psychological or psychiatric support and monitoring to identify and manage difficulties, which may arise from stress-related concerns about the diet.

#### Theme 5 – The wider context

Our findings further demonstrated the benefit of process evaluation for identifying contextual factors associated with variation in participants’ experiences and outcomes in clinical studies [16]. Across the interviews, participants discussed various influences that either supported or constrained their engagement with, or adherence to, the intervention.

##### Social pressures

Most participants commented on the interaction between their individual involvement in the study and wider social environments. Two-thirds discussed how the study influenced their family life. For many this was manageable, but for those with complex family circumstances, this presented a considerable barrier to participation.

> I’m a single parent of young children and I’m living on benefits. I’m doing voluntary work; I facilitate a self-help group…one of my children’s medical diagnosis played a role [in withdrawing from the study]. So, what happened was, staying overnight, having the brain scan and coming back, is really draining on my energy. I needed to organise childcare because I’m a single parent. It threw me slightly out of my routine. My child picked up on my stress and got quite disoriented, because we were doing things like, one day each week after school is my other child’s sports club…and this is what they’d got in their head. But this time I went via Lidl to get a particular Greek yoghurt that was important in the diet, and they just got so agitated, they nearly had a meltdown in the supermarket, because we’d gone off our normal (Participant 2).

Previous research on the ketogenic diet for children demonstrated the impact on family members, for example through nonacceptance of the diet or disruption of family routines, including mealtimes [48, 65]. Our findings extend knowledge by uncovering similar patterns within the context of adults with bipolar disorder. Notably, our study suggests that participation in the intervention may adversely affect those who have caring roles within their families (*e.g.* for dependent children or older relatives). Similar pressures, at least in the early stages of the intervention, were also identified in other social settings.

The majority of participants mentioned how being on a ketogenic diet limited their abilities to socialise in ways they were accustomed. Some recalled needing to manage questions from others about their diet.

> It was a wedding, a ladies’ evening, two graduations, so a formal dinner. And we talked about how to cope with that. And for the wedding, I took my little Pyrex dish of salad and avo[cado] and bacon…And I coped fine with it. I had to cope with questions about it but that was fine (Participant 15).

While in most cases participants adapted to the social aspect of the diet over time, felt supported to handle such situations and perceived peoples’ reactions to be *‘understanding and cool with it* (Participant 6)*,’* some found this created a talking point that raised the risk of unwanted disclosure about their condition.

> One other thing is your disclosure status, because it’s quite a visible thing to be doing, to be eating so differently. And if people don’t know your diagnosis or why you’re doing it and stuff, that is just a level of complication, why are you doing it so strictly?…I kept myself to myself quite a bit, and the people that know my situation were the people that I tended to spend more time with. But with others I just used to say I’m on a diet (Participant 6).

The challenges of eating in social settings are an everyday reality for adults following a ketogenic diet; in many cases these are temporary as people learn to manage the challenges of the diet [66]. ‘Keto-friendly’ mitigation strategies, such as bringing one’s own meals to social events, are common.

Some participants also commented on how the diet interacted with their work lives. This relationship remains under-researched.

> A lot of work commitments and things made parts of the diet difficult to do, but that was purely down to work more than anything else…the four o’clock [data] returns could be a bit tricky around work as well, but I understand why it was at that time of the day (Participant 11).

Most of these participants felt that being unemployed or able to work from home was a key factor in affording time to prepare meals and balance the requirements of the study around their other commitments. They recognised that others may be less fortunate, and there were several instances of people working away from home choosing to skip meals, struggling to find appropriate food in shops, or failing to return timely capillary blood readings.

##### Keto in a cost-of-living crisis

The pilot study took place toward the start of a period of significant economic uncertainty in the United Kingdom, characterised by high inflation and global unrest driving-up food and domestic energy prices [67]. It is notable that most participants in this study were from middle-to-higher socioeconomic backgrounds, according to SIMD data. This was reflected in many responses speculating about the potential impact of a ketogenic diet for those with less financial means. However, several participants also commented on their direct experiences of the costs associated with the intervention.

> Yes, I think it [the bills] definitely increased. I had to buy stuff for me and then different stuff for my kids. It is quite a high protein diet so that is usually a bit more expensive than carbs and then fish, fruit and veg. Things like raspberries are quite expensive (Participant 4).

Generally, the ketogenic diet is considered to be expensive for consumers compared to other diets [48, 63]. Participants recognised this as a potential barrier to those struggling with the cost-of-living, as food prices increased by 9.2% in the year to December 2022 [68]. A concurrent decrease in meat, fish and dairy sales was observed across Scotland among the general population, due to rising costs [69]. Selecting such foods appropriate for the diet in this context highlights the imperative for intensive support from ketogenic dietitians, alongside other financial-related strategies to reimburse out-of-pocket expenses as provided in the present study (e.g. travel to testing sites), to ensure inclusivity of the intervention. Mitigating the effects of a cost-of-living crisis is crucial to avoid inequalities in recruitment, adherence and health outcomes. Further economic studies are required.

#### Further suggestions for developments in future trials

Participants and research clinicians alike made numerous suggestions for developments that should be considered in related future research trials. Broadly, these related to behavioural, communication, delivery, material, psychological, social and technological factors (see Additional file 5). Some of these were addressed over the course of the pilot, for example enabling automated transfer of daily blood glucose readings to the research team [4]. The two most common suggestions from participants were as follows.

Seven of the 15 participants interviewed proposed the potential benefit of additional peer support for those living with this *‘isolating illness* (Participant 6)*.’* This may range from pastoral support for those who are *‘struggling, [to] just pick up a phone and we can have a brief chat about it* (Participant 12)*,’* to being motivationally *‘helpful as well, with making me keep at it* (Participant 4)*.’* However, despite the potential popularity of peer support, the evidence to support this in interventions for people with severe mental illness remains equivocal [70, 71]. Any such support would need to be carefully considered in the context of potential participant blinding in trials.

Linked to the challenging nature of the intervention and the societal context discussed in this paper, the same number of participants also suggested future studies may consider additional material support, especially for those facing greater financial hardship or without suitable cookery skills:

> I think the equity issue around this, and really reaching the people who, if it’s shown to work, it might benefit…And that’s not always the case for people and they don’t all have the same amount of support as I do. So I suppose for some people financial incentives might be a deal-breaker, and help buying the food or giving the food (Participant 6).

While direct financial aid and in-kind contribution, such as the provision of food parcels or pre-prepared meals on prescription [5], may support engagement among individuals from lower socioeconomic backgrounds (none of the participants interviewed were resident in the lowest SIMD quintile), people with bipolar disorder may be at greater risk of having difficulty cooking food for themselves [72]. This may be compounded by the complexities of preparing food in the ketogenic diet [5].

### Quantitative findings – fidelity checklists

Overall, there were 104 diet review meetings in which fidelity checklists were completed between participants and a research dietitian. Twenty-five of 26 participants who commenced the diet had at least one recorded meeting. Among these, 20 participants completed the diet for at least 6 weeks. Of these 20, most had five review meetings (n=15), while three participants had four (including one who declined to work through the checklist in their first meeting); two participants had three and six meetings, respectively. The remaining five participants had either one or two review meetings each.

It was intended for most, not necessarily all, behavioural components to be addressed in a single meeting, while ensuring that all were considered over the duration of the intervention. Each was addressed, as intended, at least once with 17 of the 20 participants who completed the diet for at least 6 weeks. Most components were addressed on multiple occasions with all participants. Before and after adjustment for participant withdrawal, the overall percentage of meetings that addressed a majority of components was 77%.

Figure 1 shows the percentage of diet review meetings in which the intended components were either discussed or implemented. Several behavioural components for the intervention were delivered with good fidelity. The most common component was discussing performance and providing feedback, which was documented in 93% of recorded checklists (91% non-adjusted). Setting and reviewing goals and action plans also featured prominently, perhaps reflecting its importance for people with bipolar disorder as it gives structure to their daily activities [24].Meanwhile, three behavioural components were addressed less often. Providing information to participants on the benefits of changing their diet was discussed in 42% of meetings (44% non-adjusted); 70% of all instances in which this component was raised occurred during the first two review meetings across all participants. Discussing environmental restructuring occurred in 51% of meetings, and there did not appear to be any pattern to when this component was addressed.Finally, discussing how doing well could be rewarded was addressed in 46% of meetings (42% non-adjusted). Free text comments indicated that many participants felt that this was typically an unnecessary consideration, while one dietitian stated during their process evaluation interview that they *‘sometimes found that one a little bit tricky…a little but more of a struggle to think of potentially what the reward could be* (Research Dietitian 1)*.’* This reflects previous research that suggests people find it difficult to consider non-food-based rewards for people on ketogenic diets [49]. It is proposed that participants may respond well to contingent rewards such as praise and encouragement in dietary interventions, which can support motivation and self-efficacy [73].

**Figure 1.**
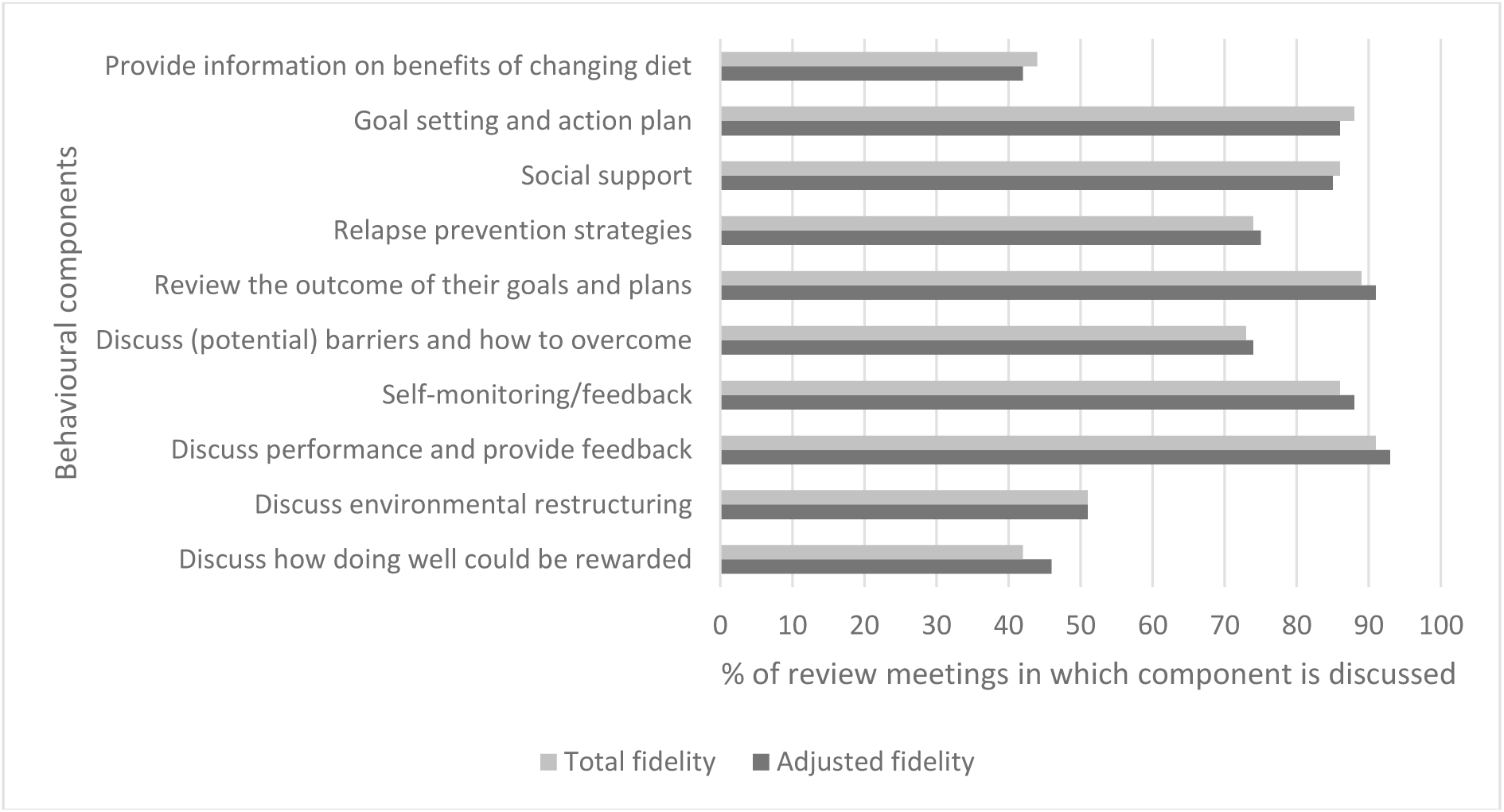
Fidelity of behavioural components implemented in diet review meetings

While many behaviour change techniques were implemented throughout the intervention period, the findings suggest that different techniques may have been used for different participants at various stages. This reflects the potential that different techniques may be more appropriate at certain times, and may better work for different people according to context and delivery method [74]. Further, different processes may need to be activated according to people’s readiness for dietary change [75]. This highlights the need to facilitate and understand participant expectations, and build rapport between participants and dietitians, for example through regular diet review meetings as in this pilot. It also indicates the importance of behavioural science support in future trials.

Overall, data indicated moderate-to-good fidelity to the behaviour change components of the intervention and have provided useful insights that can inform refinements to the behavioural strategies in future trials. Interviews with dietitians suggested that they both found the fidelity checklist a useful resource.

Because of fidelity questionnaire and because of the questionnaire that we had around things like adverse events…structured the interviews quite well. They were mostly all questions I would have asked anyway. So therefore being able to just sort of have that there, and you were recording as you were going along, then yeah, they were beneficial in that respect (Research Psychiatrist 2).

Checklists are an important strategy for examining fidelity in process evaluations and have been successfully used in dietary interventions [76, 77]. While positive experiences of using the checklist were reported in this study, there is emerging research to suggest that additional trialling of checklists at the onset of a study may improve application [78].

### Strengths and limitations of the study

This pilot study was among the first to demonstrate the feasibility, acceptability and safety of a ketogenic diet intervention for bipolar disorder [4, 15]. The process evaluation presented in this paper reinforces these observations. The intervention was theory-based and informed by current evidence regarding successful behaviour change techniques in dietary interventions. We used a rigorous multiple methods approach to data collection and analysis, and were able to triangulate data to strengthen the findings. Our research was further strengthened by the inclusion of 70% of all participants who continued the diet for 6 weeks and who completed follow-up assessments. We provide under-investigated insight into the experiences of people living with bipolar disorder initiating and following a ketogenic diet, as well as those of research clinicians who support this behavioural intervention.

However, our findings need to be considered in light of limitations in the study design and sampling. The post-hoc design of this study and short intervention period meant that findings and recommendations could not be addressed and further tested during the study. We were only able to arrange an interview with one participant who had withdrawn from the study and as such, may not have uncovered additional barriers and constraining contextual factors to engagement with a ketogenic diet. Relatedly, none of the participants in our sample were from the lowest socioeconomic areas (SIMD), where barriers may differ. Future studies should explore the potential need for additional support for these groups and others at risk of drop-out, to better understand contextual facilitators or constraints on their participation and adherence. A final limitation of our sample of research clinicians was that we only interviewed dietitians and psychiatrists. Further research is needed to understand the roles of other researchers involved in similar studies, such as imaging teams, given the potential importance of perceived positive baseline testing to the overall participant experience (*i.e.* feeling informed, cared for, and generally at ease). Additional areas of future work include further exploration of the motivation and engagement of the family in relation to providing social support, and mechanisms of dietary change.

## Conclusions

The findings provide valuable insight into the process of delivering and engaging in a ketogenic diet intervention for people living with bipolar disorder. In doing so, we highlight how challenging the ketogenic diet can be for some participants. However, for most people the intervention was considered feasible and acceptable and was delivered with good fidelity. Many participants reported a range of physical and psychological benefits, despite common side-effects, some less-positive experiences and some disruption to elements of daily and social living. To offset or overcome barriers such as dietary preferences, concerns about the diet and its possible impact, administrative burdens and limited implementation capacity, we advocate the incorporation of varied behavioural and social support strategies and technology assisted monitoring. Future trials may benefit from increased researcher/research team capacity, better-defined intervention entry and exit routes, additional interpersonal support, and a greater understanding of how social and societal factors impact participation among people with bipolar disorder, particularly among those most likely to be affected by these.

## Data Availability

Data generated and analysed during the current study are not publicly available as explicit consent was not sought from participants, and privacy may be compromised.

## Electronic supplementary material

Additional file 1: Interview topic guide (pilot participants)

Additional file 2: Interview topic guide (research clinicians)

Additional file 3: Intervention fidelity checklist

Additional file 4: Perceived benefits and down-sides of the ketogenic diet reported during interviews

Additional file 5: Suggestions for developments in future trials

## List of abbreviations

BMI: Body mass index
RCT: Randomised controlled trial
SIMD: Scottish Index of Multiple Deprivation

## Declarations

### Ethics approval and consent to participate

The study received a favourable ethical opinion from the South East Scotland Research Ethics Committee 02 (approval number: 22/SS/0007) and was approved by NHS Lothian Research and Development (2022/0009). Sponsorship was provided by the Academic and Clinical Central Office for Research and Development. Written informed consent was obtained from all participants. The study was prospectively registered in the ISRCTN Registry under registration number ISRCTN61613198 on 2 March 2022.

### Consent for publication

Not applicable.

## Funding

This study was funded by the Baszucki Brain Research Fund. BPR and SAS acknowledge funding from the UKRI Medical Research Council (MC_UU_00022/1) and the Chief Scientist Office of the Scottish Government Health Directorates (SPHSU16). MJT acknowledges funding from the Chief Scientist Office of the Scottish Government Health Directorates through the NHS Lothian Research and Development Office.

### Authors’ contributions

All authors contributed to the conceptualisation and design of the process evaluation. NN, HG and BPR were responsible for sampling and recruitment. HG and CF collected the fidelity checklist data; NN, HG, IK were responsible for the preparation of checklist fidelity data for analysis. BPR was responsible for interview data collection. BPR, SAS were responsible for analysis of the data. Data were interpreted by BPR, SAS (qualitative data) and BPR, SAS, NN, HG, IK (quantitative data), and these interpretations were discussed among all authors through team meetings. BPR was responsible for drafting the paper. All authors were involved in subsequent revisions and agree to be accountable for all aspects of the work.

## Acknowledgements

We the authors wish to thank all participants and research clinicians who were involved in this research project.

## Additional file 1: Interview topic guide (pilot participants)

### Preamble

Today, I am going to ask you to tell us about your experiences of being involved in our study. I will ask you about your experiences of the ketogenic diet, the support you received, and how you felt about being a research participant. There are no right or wrong answers. Feel free to say as much or as little as you like, and remember that you do not have to provide an answer to a question if you do not want to. I may ask you to discuss things in a bit more detail, but if you do not feel you have anything further to say, that is fine. Some of the things we discuss today may be sensitive, and everything will be treated in confidence. We can take a break at any time, just let me know. If you want to finish the interview early, please say so, and be assured that you do not need to provide a reason for this.

Are you happy for me to start the recording?

### Background and context

- Can you tell me how long you have been living with bipolar disorder and how it has impacted you?

### Taking part in the research: recruitment

- How did you find out about the study? What attracted you toward it?
- What information did you receive about the study? Was it helpful/what could be improved?
- Why did you decide to participate? Did you have any initial concerns?

### Taking part in research: your experience

- How manageable was being a participant in this study?
- Did you face any barriers to taking part as in the study?
- Were the expectations on your time realistic? How did the study fit around your existing daily life? How did it impact those around you (*e.g.* at home or at work)?
- Did you feel able to keep up with the testing requirements (*e.g.* completing diaries or remembering to do blood tests)?
- Can you tell me a bit about the hospital visits – How was attending for MRI scans and venous blood tests before the study and at follow-up?
- I would like you to tell me a little bit about the different Apps and questionnaires that we used in this study (*e.g.* were they easy to use/not; can anything make them more useful):
- How did you find completing the mental health and health economics questionnaires before the study and at the follow-up? What did you think about being text daily for data (if used)?
- What did you think about using the Ketomojo and ilumivu apps (if used)?
- What did you think about wearing an Actigraph device on your wrist?
- Throughout this study your diet and health were very closely monitored, how did this make you feel?
- How did taking part in this study interact with any ongoing management or treatment that you have for bipolar disorder?
- How could we better incentivise participants to take part in a study like this?

### The diet

- What came to mind when you thought about making changes to your diet? Did the diet meet your initial expectations?
- What were the easier/harder aspects of the diet to put in place?
- How well did it fit into your existing eating patterns (*e.g.* were you able to retain the same mealtimes/did you find yourself snacking more)?
- Was there anything that you did to help you ‘stick to’ the diet? How was this helpful/not?
- What did you like and dislike about the ketogenic diet?

### Support package

- During the study, what type of help (info/resources/support) was the most helpful in relation to the diet? What was least helpful? Why?
- What do you think about the individualised recipe leaflets that you were provided?
- Did you use the keto-calculator? If so, what did you think about this?
- Thinking about the research dietitian…
- Can you tell me a little bit about the initial meeting, how did that go?
- What support did you receive from them regarding the diet? Did you ask for anything specific?
- How did you choose to interact with them (*e.g.* telephone, email, face-to-face)? Do you have a preference, and if so, why? How much contact time do you think is about right?
- How did the support you received compare to the expectations that you had at the start of the diet?
- Thinking about the study coordinator:
- Did you engage with them? If so, are you happy to tell me about that experience?
- What support did you receive from them regarding the study and diet?
- How much contact time with the study team do you think is about right?

### Outcomes

- Has the study helped you? How/why not?
- How have things changed for you as a result of the study? Have you seen changes in other parts of your life apart from diet?
- Did you experience any negative side-effects from the diet? How did you manage these? How long did they last, were they tolerable?

### General

- What was the most important positive aspect of this experience for you?
- Is there anything important that you would recommend that we change about i) the study; or ii) the ketogenic diet intervention, if we were to do it again?
- Do you have any further thoughts or reflections about this study?

## Additional file 2: Interview topic guide (research clinicians)

Today, I am going to ask you to speak about your experiences of being involved in the study as a research clinician. I will ask you about your experiences of implementing the intervention and supporting participants. There are no right or wrong answers, and you should feel free to say as much or as little as you like. You do not have to provide an answer to a question if you do not want to. I may ask you to expand on your answers at times, but if you do not feel you have anything further to say, that is fine. We can take a break at any time, just let me know. Everything we discuss will be treated in confidence. If you want to finish the interview at any time, you can do so without needing to provide a reason for this.

Are you happy for me to start the recording?

### General experiences

- Can you tell me a bit about your experience of working on this study? How did you find it?

### Implementing the study

- Did you find the study paperwork easy to complete? If not, what issues were there?
- What were the barriers/facilitators for you to implementing the intervention? How did you try to overcome any barriers that you faced?
- What was your experience of using the Apps and other monitoring tools? Did you feel suitably equipped to support participants with these?
- Considering the fidelity checklist…was this useful?
- Which components of the intervention (including the behavioural strategies) did you implement more or less often?
- How did you try to incorporate the different behavioural strategies? Were any particularly easy/challenging to use?
- Were there aspects of the study that you struggled to implement as intended? If so, why?
- Did you make any changes to the way the intervention was delivered from the protocol?
- How did delivering the intervention and the different study components correspond to your usual working practices? Was it challenging?

### Participants

- Through what media did the participants engage with you? What were the kinds of reasons they would get in touch?
- From your perspective what seemed to be the barriers/facilitators to participant engagement with the diet?

### Moving forward

- If we were to conduct this study in future…
- What additional support would research clinicians require?
- Do you have any suggestions as to how you might improve compliance with the diet?
- Do you have suggestions as to how we might increase recruitment into the study?
- Is there anything important that you would recommend that we change about the study or the intervention if we were to do it again, and why?
- Do you have any further thoughts or reflections about this study?

## Additional file 3: Intervention fidelity checklist

| Element | 1 | 0 | Comments |
| --- | --- | --- | --- |
| Provide information on benefits of changing diet (e.g. the medical and personal benefits, thinking of the intervention not as a diet but a medication/treatment). |  |  |  |
| Goal setting and a plan of action: (i.e. set or discuss/review goals to change behaviour). |  |  |  |
| Social support (e.g. identify relevant family/friends who can offer support; other people also on keto diet e.g. via cooking classes) |  |  |  |
| Relapse prevention strategies (e.g. ways to avoid eating certain foods; not putting themselves in certain situations) |  |  |  |
| Review the outcome of their goals and plans (i.e. were the patient's achievement of their goals/progress discussed). |  |  |  |
| Discuss (potential) barriers and how to overcome these (e.g. side effects, potential negative influence of family members). |  |  |  |
| Self-monitoring/feedback (e.g. monitoring blood every day; monitoring mood). |  |  |  |
| Discuss participant's performance (including success/failure) and provide feedback. |  |  |  |
| Discuss environmental restructuring options (e.g. get rid of biscuits in the cupboard). |  |  |  |
| Discuss how doing well could be rewarded (e.g. a trip to the cinema or other treat). |  |  |  |
| General comments. |  |  |  |

1 = element conducted; 0 = element not conducted

## Additional file 4: Perceived benefits and down-sides of the ketogenic diet reported during interviews

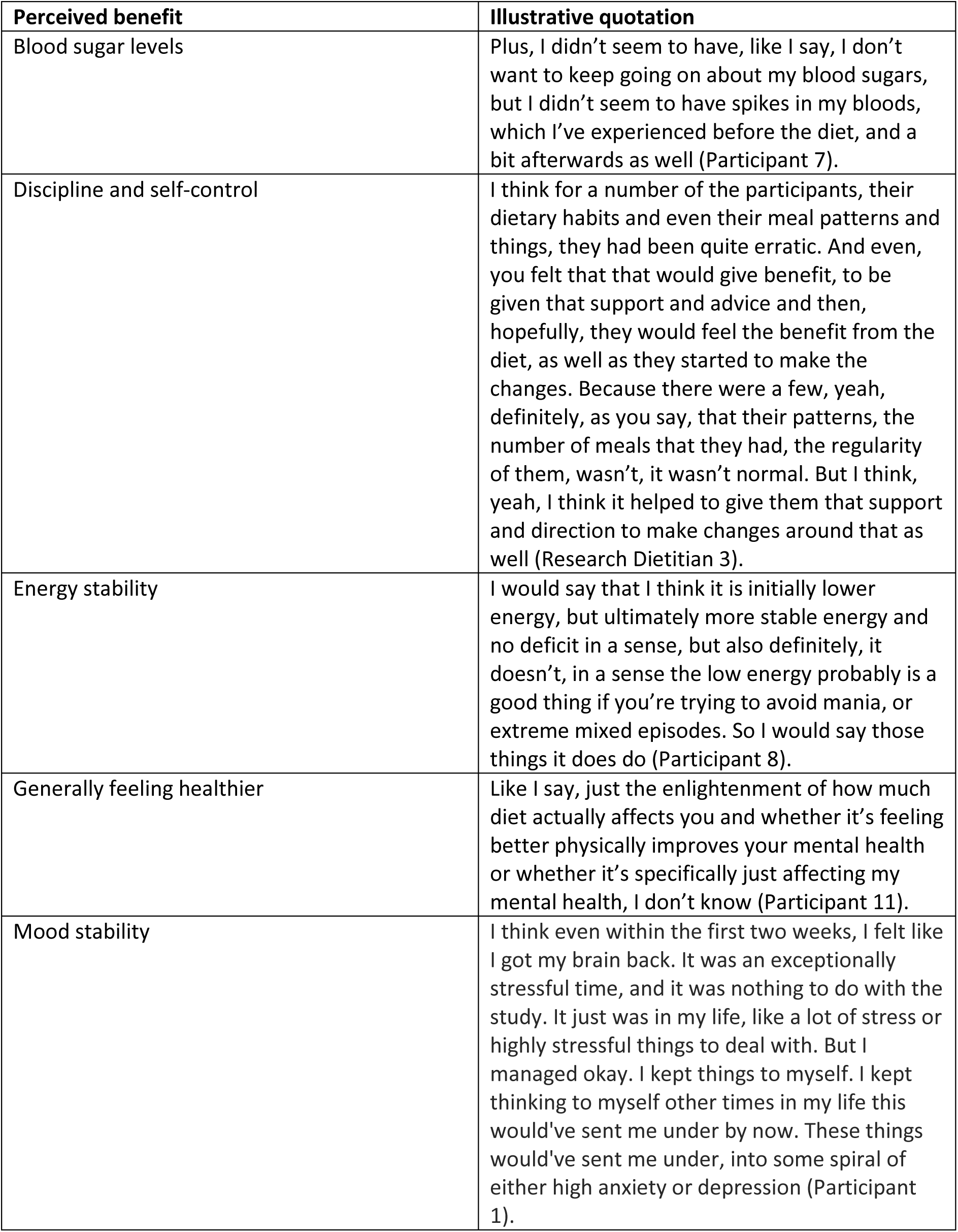
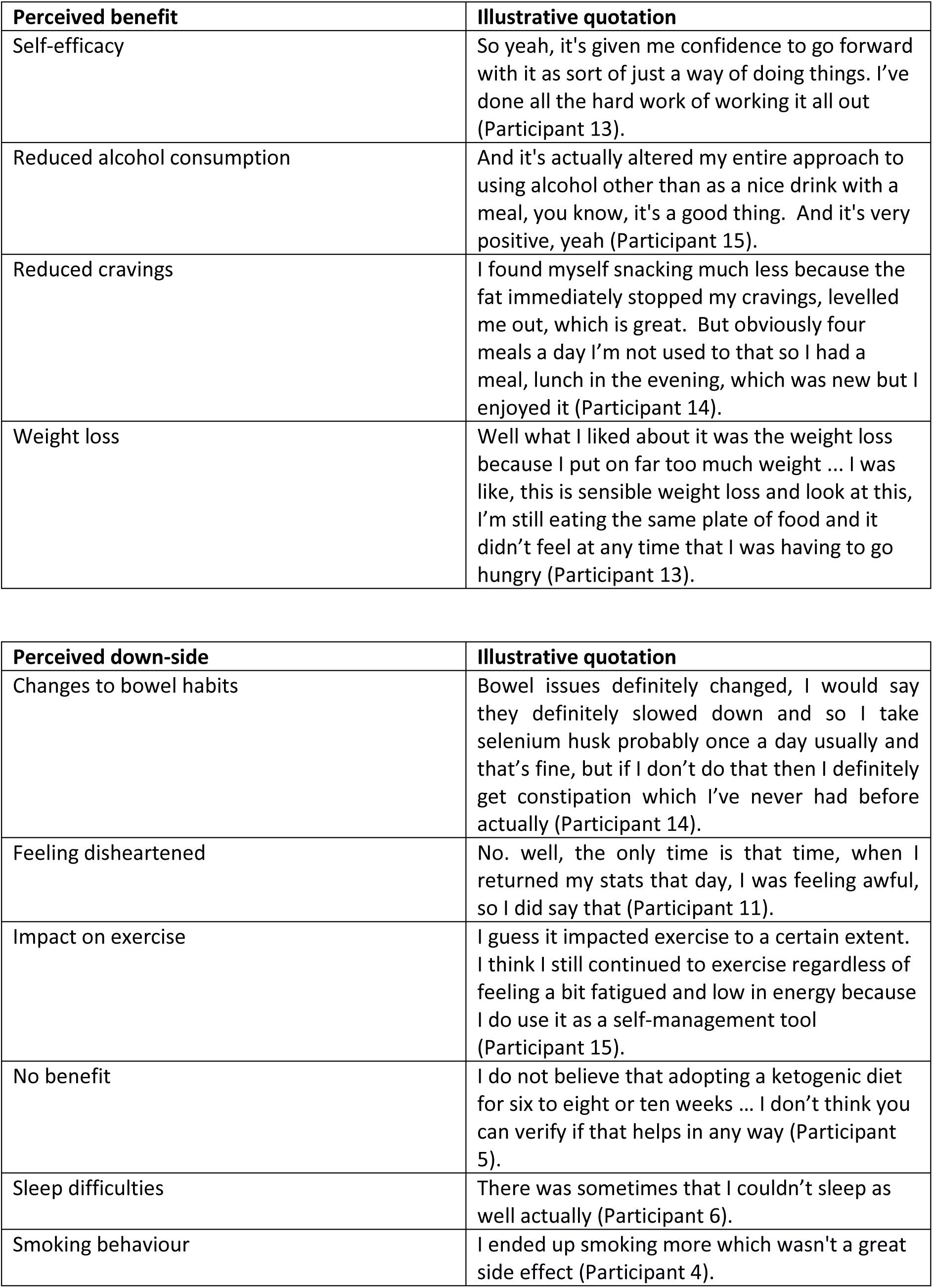

## Additional file 5: Suggestions for developments in future trials

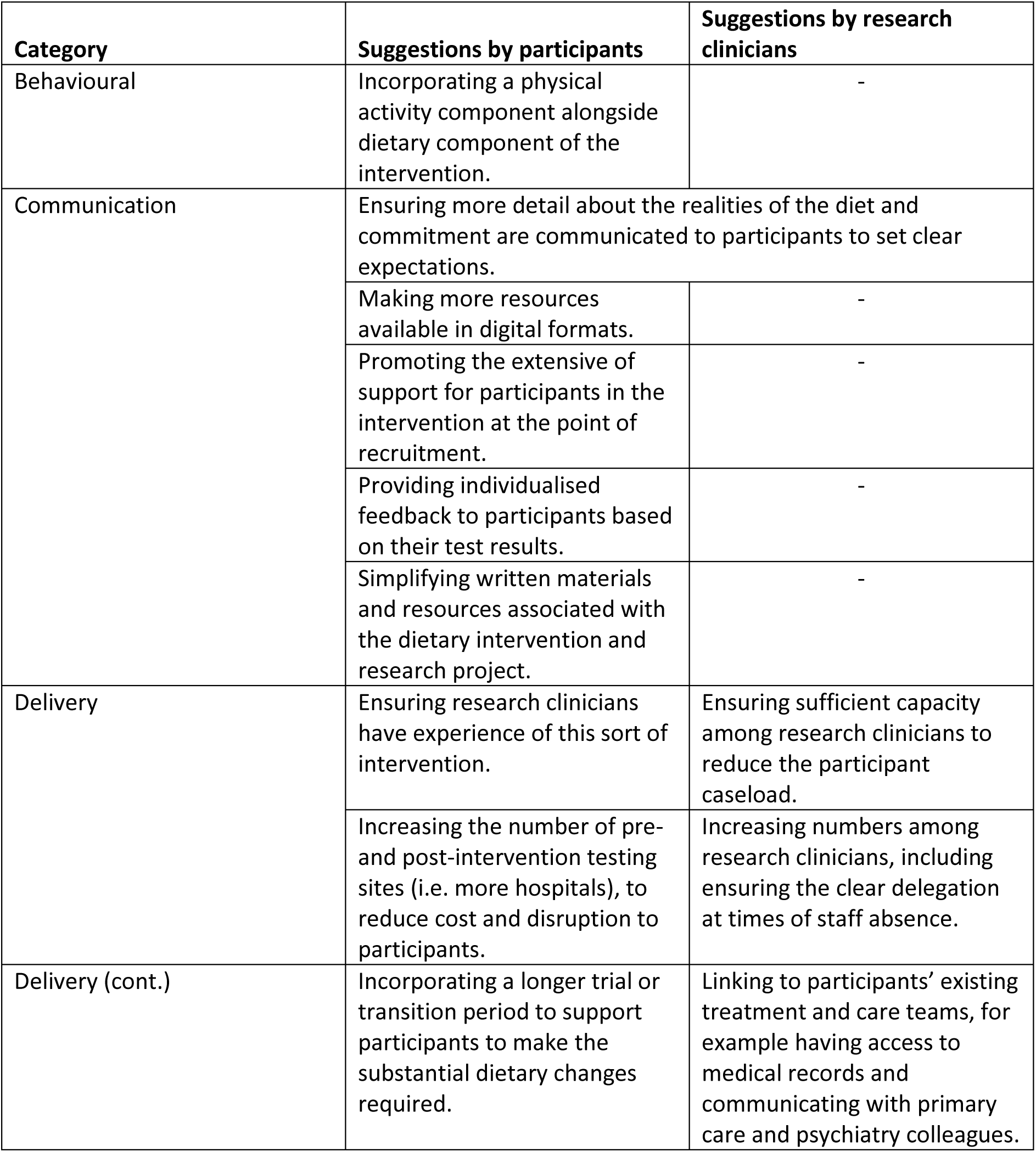
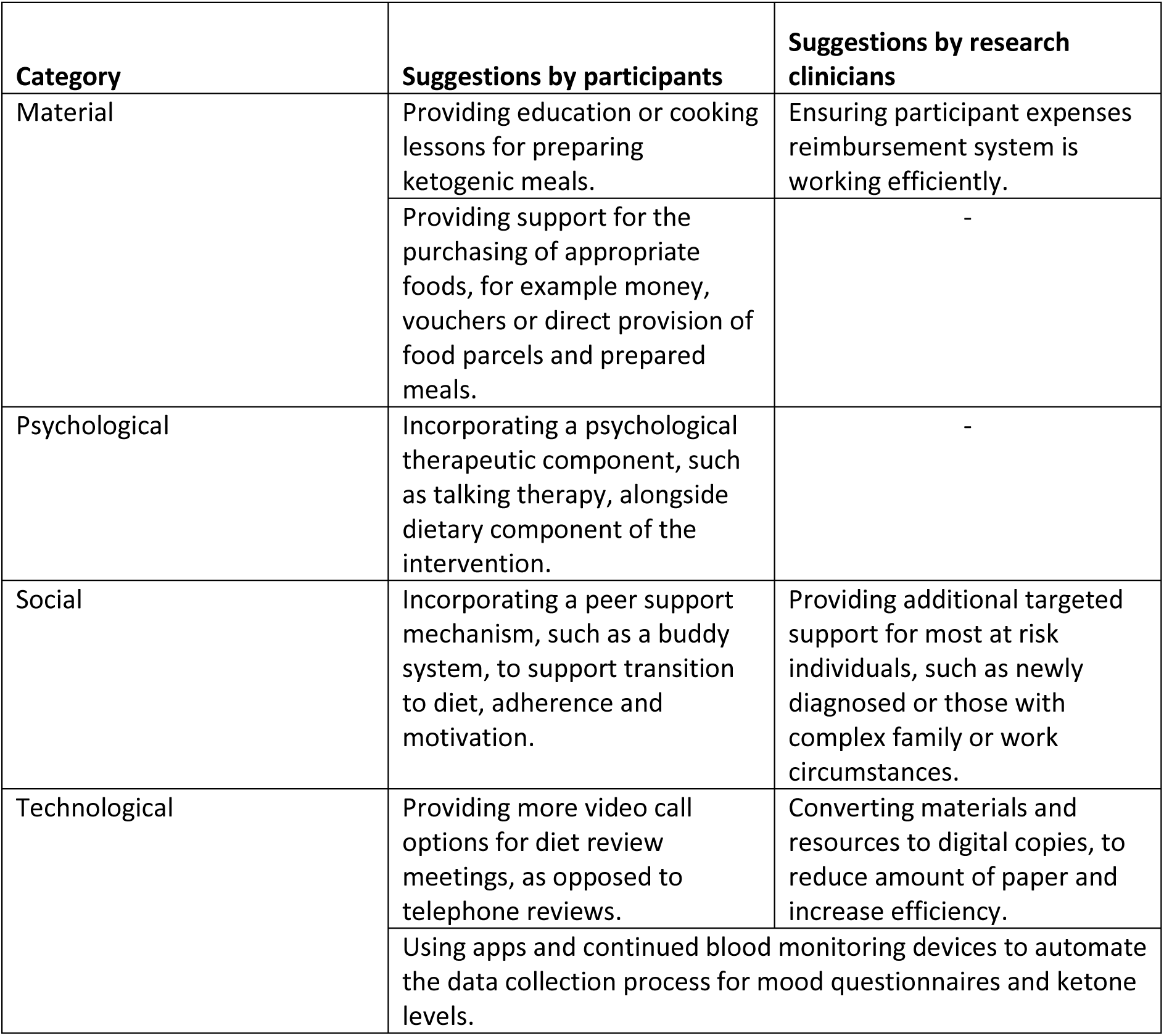

| Category | Suggestions by participants | Suggestions by research clinicians |
| --- | --- | --- |
| Behavioural | Incorporating a physical activity component alongside dietary component of the intervention. | - |
| Communication | Ensuring more detail about the realities of the diet and commitment are communicated to participants to set clear expectations. |  |
|  | Making more resources available in digital formats. | - |
|  | Promoting the extensive of support for participants in the intervention at the point of recruitment. | - |
|  | Providing individualised feedback to participants based on their test results. | - |
|  | Simplifying written materials and resources associated with the dietary intervention and research project. | - |
| Delivery | Ensuring research clinicians have experience of this sort of intervention. | Ensuring sufficient capacity among research clinicians to reduce the participant caseload. |
|  | Increasing the number of pre- and post-intervention testing sites (i.e. more hospitals), to reduce cost and disruption to participants. | Increasing numbers among research clinicians, including ensuring the clear delegation at times of staff absence. |
| Delivery (cont.) | Incorporating a longer trial or transition period to support participants to make the substantial dietary changes required. | Linking to participants' existing treatment and care teams, for example having access to medical records and communicating with primary care and psychiatry colleagues. |
| Material | Providing education or cooking lessons for preparing ketogenic meals. | Ensuring participant expenses reimbursement system is working efficiently. |
|  | Providing support for the purchasing of appropriate foods, for example money, vouchers or direct provision of food parcels and prepared meals. | - |
| Psychological | Incorporating a psychological therapeutic component, such as talking therapy, alongside dietary component of the intervention. | - |
| Social | Incorporating a peer support mechanism, such as a buddy system, to support transition to diet, adherence and motivation. | Providing additional targeted support for most at risk individuals, such as newly diagnosed or those with complex family or work circumstances. |
| Technological | Providing more video call options for diet review meetings, as opposed to telephone reviews. | Converting materials and resources to digital copies, to reduce amount of paper and increase efficiency. |
|  | Using apps and continued blood monitoring devices to automate the data collection process for mood questionnaires and ketone levels. |  |

